# Effect of Cardiac Fiber Orientation on Electrical Dyssynchrony in Ventricular Ectopy

**DOI:** 10.1101/2025.07.12.25331148

**Authors:** Sidney James Perkins, Matteo Salvador, Zinan Hu, Oguz Tikenogullari, Fanwei Kong, Sanjiv Narayan, Alison Marsden

## Abstract

**Background:** Widened QRS complex confers worse prognosis in both pacing and premature ventricular contraction (PVC), and may predispose to heart failure. In healthy adult patients, cardiac fiber orientation is highly variable. While traversing the myocardial wall from epicardium to endocardium, helix angles are known to range between -90° and 90°. It is currently not well understood how the location of ectopy and variable epicardial and endocardial helix angle configurations interact to impact the underlying QRS duration of ectopic beats. Methods for identifying regions of favorable pacing accounting for individualized fiber orientation and cardiac geometry are currently lacking. In this work, we hypothesize that fiber orientation will impact QRS duration for pacing locations along the right ventricular (RV) septum. Computational modeling provided an efficient platform in which to systematically study these effects in a controlled manner.

**Methods:** Five fiber phenotypes with endocardial and epicardial helix angles ranging from 40° to 80° and -80° to -40° were studied, respectively. Three control pacing conditions were considered, using a fractal tree-generated Purkinje network, and a model of right and left bundle branch block (RBBB, LBBB). Five pacing locations along the inferior RV septum were considered. The monodomain equation coupled with the ten Tusscher Panfilov ionic model was used in finite element simulations of electrical propagation. Anisotropic and isotropic conduction velocities were 0.49 m/s and 0.21 m/s, respectively. Electrocardiograms (EKGs) were generated for each condition, and the QRS duration was measured as the variable of interest.

**Results:** Variable fiber phenotype prolonged the QRS duration by 1.88, 4.49, and 7.48 ms (3.92, 2.35, and 4.70%) in the Purkinje, RBBB, and LBBB conditions. In five pacing locations along the inferior RV septum, QRS duration prolonged by between 14.33 and 27.00 ms (6.72 and 13.02%). In addition, the site associated with the shortest and longest QRS duration differed depending on the fiber phenotype.

**Conclusions:** We introduce a pipeline to study individualized impacts of cardiac geometry and fiber orientation on QRS duration for variable pacing conditions. Our findings implicate fiber orientation as a risk factor for development of cardiomyopathy and suggest a possible role of preoperative fiber orientation characterization prior to pacemaker placement.

## INTRODUCTION

In normal cardiac physiology, pacemaker cells in the sinoatrial node initiate cardiac contraction and set the intrinsic heartrate. Depolarization then proceeds inferiorly through the atria, atrioventricular node, His-Purkinje system, and terminates with ventricular depolarization. Occasionally, the cardiac ventricles contract prematurely, which can result in an uncomfortable sensation. Premature ventricular contractions (PVCs) are common, present in roughly 6% of patients with 2-minute electrocardiograms (EKGs).(1) Typically PVCs are thought to be benign and carry an excellent prognosis, however, roughly 30 - 40% of patients with PVC burden greater than 10% may develop a reversible form of heart failure known as PVC-induced cardiomyopathy.(2)

Catheter ablation is the first-line therapy for PVC-induced cardiomyopathy.(3) However, because there is a limited understanding of the fundamental pathophysiology of PVC-induced cardiomyopathy, it is difficult to determine which patients will progress to cardiomyopathy and benefit most from PVC ablation prior to the onset of heart failure signs and symptoms. While the exact pathophysiology is poorly understood, known risk factors for development of PVC myopathy include increased PVC burden, longer QRS-complex duration, epicardial origin of PVCs and asymptomatic PVCs.(3–5) Porcine models in PVC myopathy have pointed toward mechanical dyssynchrony as a potential culprit. In one study considering right ventricular paced bigeminy in a sample of ten pigs, the magnitude of decreased ejection fraction was noted to correlate strongly with the percent of ectopic beat dyssynchrony. (6)

Like heart failure arising in the context of increased burden of PVCs, patients with pacemakers are at an increased risk of developing myopathy. In a prospective cohort study following 363 patients undergoing dual-chamber and single-chamber ventricular pacemaker implantation, roughly 13.8% of patients were noted to develop pacemaker-induced cardiomyopathy (PICM) over a mean follow-up of 14.5 months. (7) Similar to what is seen in PVC cardiomyopathy, those patients who had a higher burden of right ventricular pacing developed myopathy more frequently (HR 4.26 *p*=0.004). Moreover, similar to what was shown in the porcine model of PVC myopathy, those patients with increased mechanical dyssynchrony developed heart failure at a higher rate than those without mechanical dyssynchrony (HR 3.15 *p*=0.002). (7)

Taking together what is currently known about prognosis in pacemaker-induced and premature ventricular contraction cardiomyopathies, there is compelling evidence that electromechanical dyssynchrony between the left and right ventricles confers a greater risk of developing cardiomyopathy.

Still, mechanistic connections between risk factors and increased risk of developing electromechanical dyssynchrony for any given anatomical location of ectopy remain unknown. Two patients with the same burden of ectopic beats with the same origin of ectopy may have different clinical outcomes. One potential factor that may stratify risk for developing electromechanical dyssynchrony in patients with both PICM and PVC myopathy is cardiac fiber orientation. Though fiber orientation is not routinely available on diagnostic cardiac MRI (CMR), specialized experimental sequences obtained through diffusion tensor MRI (DT-MRI) are able to provide information about region-specific cardiac fiber orientation and are of increasing interest.(8)

Cardiac fiber orientations change gradually as the myocardial wall is traversed from epicardium to endocardium. Many groups have used DT-MRI to quantify human cardiac fiber orientation in vivo and ex vivo in otherwise healthy subjects, noting a large variation in epicardial and endocardial helix angle. A study of ten healthy volunteers by Nielles-Vallespin, et al. used DT-MRI *in vivo* and reported helix angles of -17 ± 7° and 23 ± 9° (mean ± STD) epicardially and endocardially, respectively.(9) Others have found a wider distribution of fiber orientations for healthy volunteers. Tseng, et al measured fiber orientations in five healthy volunteers (three male, two female) ages 26-41 years old, and reported helix angles for only four subjects. Their figures demonstrate helix angles of -30 ± 22° and 63 ± 28°, epicardially and endocardially, respectively.(10) Toussaint, et al studied five healthy volunteers and reported a range of fiber orientations between -77° and 0° epicardially and 46° and 83° endocardially.(11) Finally, similar methods have been used *ex vivo* for otherwise healthy but recently deceased humans. The advantage of this approach is that cardiac tissue can be fixed, and accurate measurements can be made without the need for cardiopulmonary gating. The largest study of this nature was performed by Lombaert, et al in 2011 and was comprised of ten individuals between the ages of 17 and 74 years (eight male, two female). They reported helix angles of -41 ± 26° and 66 ± 15°, epicardially and endocardially, respectively.(12)

Computational methods are well suited to appraise the effect of varied fiber phenotype on electrical dyssynchrony in a model of PVC myopathy and PICM. Computational methods in electrophysiology are of increasing interest for the creation of patient specific models that non-invasively address clinical and translational problems. Foundational computational work in cellular electrophysiology has produced phenomenological models to simplify and recapitulate the results of the analytic models described first by Hodgkin-Huxley in 1952.(13–16) More contemporary work has described biophysical models, that account for subcellular ion channels and have improved accuracy.(17–21) One of the most widely used biophysical models is the ten Tusscher-Panfilov model, which has been widely used to study spiral wave propagation, cardiac fibrosis, and ventricular tachycardia.(18,22,23) In addition, the ten Tusscher-Panfilov model has been coupled with lumped-parameter models to simulate the cardiac cycle and hemodynamics in both physiologic and pathologic settings.(22,24) Software such as openCARP and MyoKit have made cardiac electrophysiology modeling more accessible, and have implemented versions of commonly used models such as the ten Tusscher-Panfilov model.(25,26) Finally, due to variations in anisotropic and isotropic conduction velocities, several groups have also established methods for prescribing endocardial and epicardial helix angles for use in conjunction with biophysical models of cardiac depolarization.(27–29)

Developing a deeper understanding of the pathophysiology of PVC myopathy and PICM would enrich the field of electrophysiology, and offer clinicians valuable new tools for diagnosis, prognostication, and treatment. In the case of PVC myopathy, knowledge of a patient-specific risk of developing cardiomyopathy could provide an innovative way to risk-stratify patients undergoing cardiac ablation. In the case of pacemaker implantation, knowledge of the risk conferred by a particular location of ectopy in a patient-specific case would guide electrophysiologists in optimizing patient-specific lead placement. The objective of this study is therefore to elucidate the pathophysiology of PVC myopathy in a virtual patient dataset generated by systematically varying parameters in the setting of computational modeling. Specifically, we hypothesize that fiber orientation will alter QRS duration for variable pacing locations along the right ventricular septum. In addition, we hypothesize that the pacing location corresponding to the shortest QRS will differ depending on the underlying cardiac fiber phenotype.

## METHODS

### Patient Data and Mesh

A coronary CT angiogram of a healthy male patient was used to generate a tetrahedral 3D cardiac mesh with average edge length of 0.5 mm comprising 14,224,211 elements. Image segmentation was performed in SimVascular to produce a patient-specific biventricular anatomic model. Post-processing to define surfaces associated with the volume mesh was performed in custom Python script.

### Modeling Overview

Simulations were performed in svMultiPhysics, a C++ high-performance computing multi-physics and multiscale finite element solver for cardiac and cardiovascular electro-mechanical modeling, made available open source as part of the SimVascular project. We employ the Monodomain equation coupled with the ten Tusscher-Panfilov ionic model.(17,30) Linear tetrahedral elements were used together with a generalized-alpha method for time-discretization. Five pacing locations were chosen along the inferior right ventricular septum. Normal conduction was simulated using a fractal-generated Purkinje fiber network.(31) Right and left bundle branch blocks were simulated by omitting either the right or left Purkinje fiber network. A rule-based method implemented in Python v3 was used to generate fiber directions from prescribed epicardial and endocardial helix angles.(27) Five fiber orientation conditions were generated to characterize the parameter space described in the DT-MRI literature.

### Electrophysiology Parameter Tuning

A tetrahedral mesh with edge length of 0.5 mm was generated to represent a slab of cardiac tissue measuring 3 mm x 7 mm x 20 mm and comprising 51,879 elements. This mesh was used to study the effects on conduction velocity in conductivity tuning experiments. A pacemaker region measuring 1.5 mm x 1.5 mm x 1.5 mm in the corner of the slab was held at - 35.714 mV for 2.0 ms to initiate signal propagation. Simulations were run for 2,000 timesteps with a timestep size of 0.10 ms. In general, anisotropic conductivity was swept from 3.98*10^-9^ m^3^ / s to 2.00*10^-7^ m^3^ / s. Fiber direction was made parallel to the longest dimension of the mesh. Each conductivity experiment was performed at anisotropic to isotropic conductivity ratios of 3:1, 5:1, 7:1, and 10:1. The time associated with depolarization of the node furthest from the pacing site in the fiber and sheet directions was measured for each condition and used to compute conduction velocity. For each conductivity ratio, the relationship between anisotropic conductivity and anisotropic conduction velocity was characterized using a second order polynomial in Prism v10. In addition, the relationship between anisotropic conductivity and isotropic conduction velocity was similarly characterized. Each resulting model relating anisotropic conductivity to conduction velocity is reported as well as the R-squared values.

An anisotropic conduction velocity of 0.5 m/s with a 3:1 anisotropic to isotropic conduction velocity ratio was desired.(32) To approximate these ideal values, the results of the conductivity tuning study were used to select conductivities of 7.025*10^-8^ m^3^ / s and 7.025*10^-9^ m^3^ / s for anisotropic and isotropic conductivities, respectively. This final condition was used in subsequent patient-specific electrophysiological modeling, and the associated velocities obtained using the benchmark mesh are reported.

### Fiber Orientation Selection

A literature review was performed to identify studies characterizing the endocardial and epicardial helix angle using cardiac DT-MRI in normal adults. The authors, year of publication, species, sample size, age range, sex, and helix angles were all noted. A combination of average helix angle, standard deviation, and range of helix angles are reported, depending on statistics reported in the primary source. In some cases, only regional measures of helix angle were reported, so a global measurement was computed using the law of total variance assuming equal size and large sample sizes when appropriate.(33,34) Several papers provided figures but no summary data. For these, ImageJ v2.0.0-rc-69 and MATLAB R2024A were used to extract summary statistics.(10,11,35) Using a rule-based algorithm for assigning myocardial helix angle to a patient mesh, a total of five meshes were generated with endocardial helix angles ranging from 40° to 80° and with epicardial helix angles ranging from -80° to -40°.(27) Biventricular models, including helix angle data, were visualized using Paraview v5.10.0-RC1.

### Electrocardiogram Definition and Pacing Conditions

A twelve-lead electrocardiogram (EKG) was digitally produced for each simulation by placing virtual leads as shown in **Fig. 1(a)**. Ectopy was modeled by pacing from a series of locations along the inferior right ventricular septum (**Fig. 1(b)**). Normal conduction was modeled using a fractal-generated Purkinje network. Bundle branch blocks were modeled by selectively loading either the right or the left Purkinje network (**Fig. 1(c)**).

**Fig 1.**
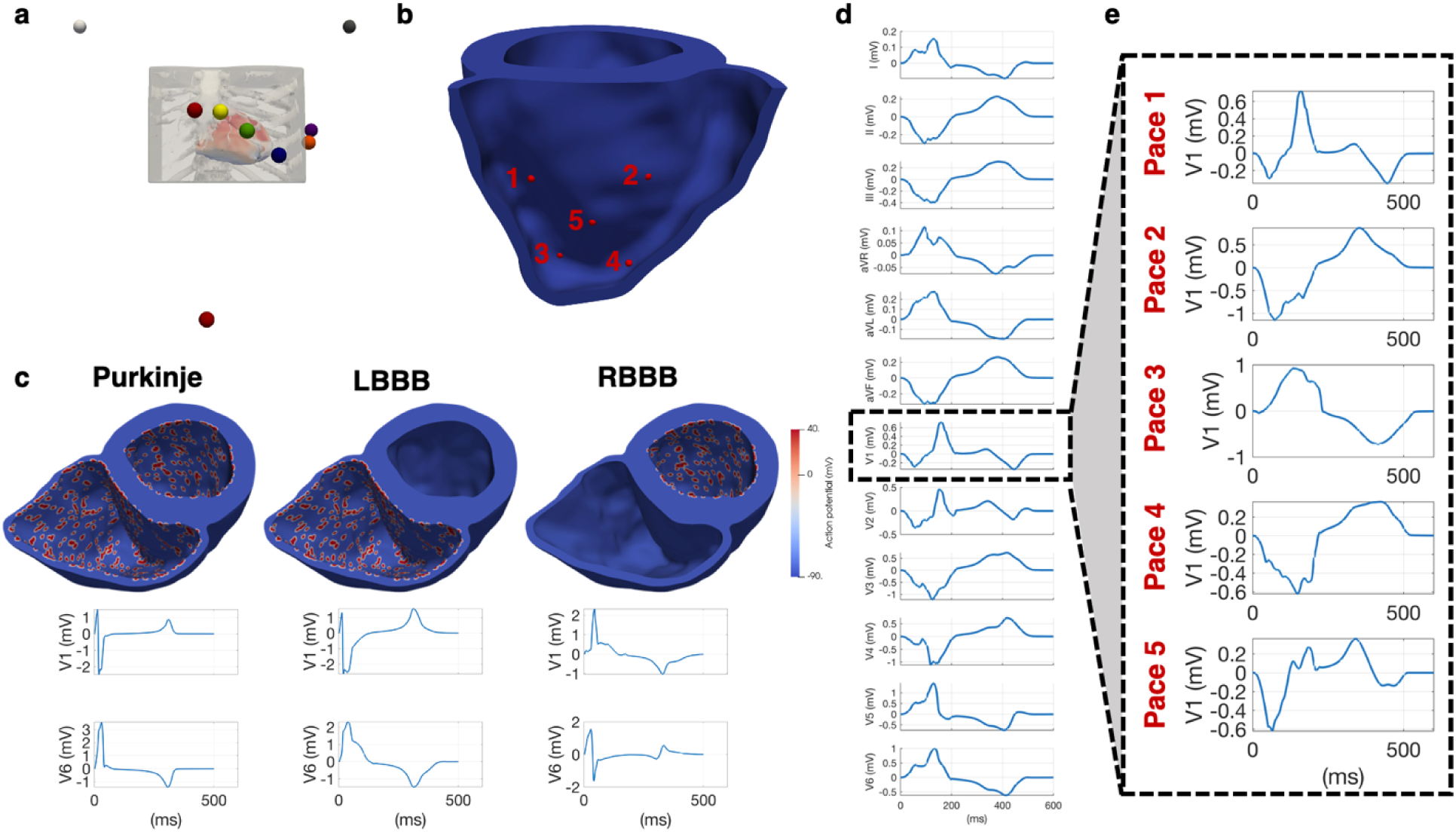
(a) Lead locations for generation of twelve-lead electrocardiogram (EKG). (b) Five sites of ectopy were considered along the inferior right ventricular septum. (c) Simulations were performed on a mesh with epicardial helix angle = -40° and endocardial helix angle = 40°. V1 and V6 are shown for Purkinje, left bundle branch block (LBBB), and right bundle branch block (RBBB). In each of the bundle branch block cases, the QRS is widened. In LBBB, V6 is positive and V1 is negative toward the end of the QRS; in RBBB, the opposite is observed. (d) A twelve lead EKG is shown for the simulation. (e) The effect of altered pacing location is shown.

### Effect of Fiber Orientation on Depolarization Dynamics

For each of the five fiber orientations and each of the eight pacing conditions, a twelve-lead EKG and activation map were created. The QRS corresponding to each of twelve leads was measured, and the maximum was used to represent the QRS for a given experimental condition. QRS widths were manually measured five times. For the normal Purkinje and the right and left bundle branch conditions, an ANOVA comparing the QRS durations associated with each fiber orientation was performed. Follow-up t-testing was performed comparing the highest and lowest QRS values within each pacing condition. Additionally, for each fiber orientation, an ANOVA comparing the QRS durations associated with each of the five ectopy locations was performed. Similarly, follow-up t-testing was performed comparing the highest and lowest QRS values within each fiber orientation condition. Significance was determined by α = 0.01.

## RESULTS

### Electrophysiology Parameter Tuning

The results of the electrophysiology parameter tuning performed on a well-defined geometry are shown in **Fig 2**. The conductivities used in the patient-specific modeling portion of this study were 7.025*10^-8^ m^3^ / s and 7.025*10^-9^ m^3^ / s for anisotropic and isotropic conductivities, respectively. These resulted in an anisotropic conduction velocity of 0.49 m/s and isotropic conduction velocity of 0.21 m/s.

**Fig 2.**
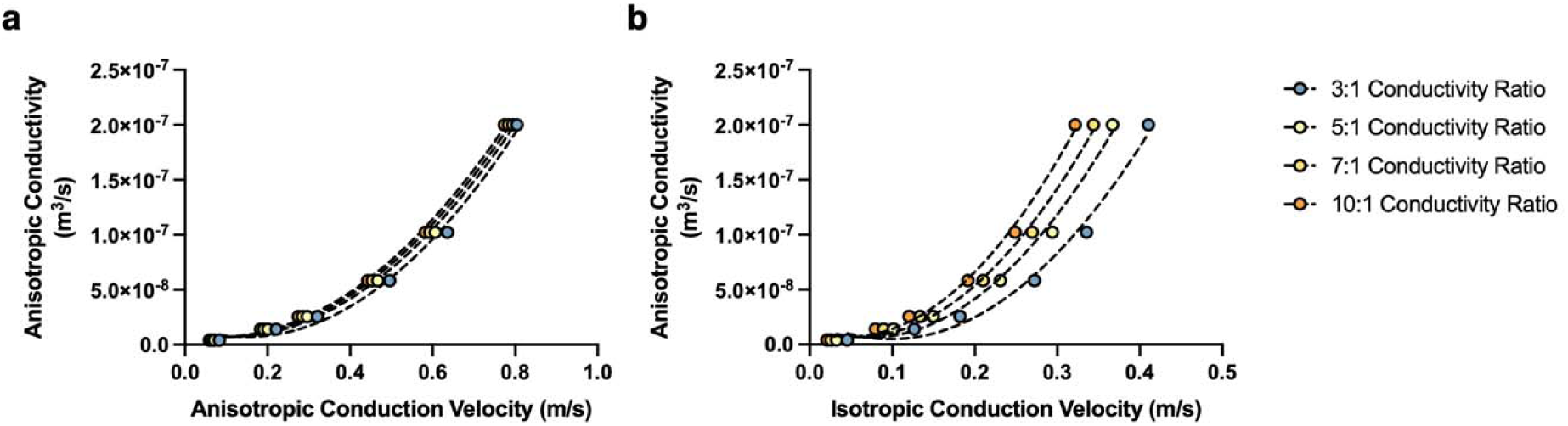
(a) The relationship between anisotropic conduction velocity and anisotropic conductivity was characterized. For a 3:1 anisotropic to isotropic conductivity ratio, the relationship between anisotropic conduction velocity (V_ani_) and anisotropic conductivity (C_ani_) was C_ani_ = 4.37*10^-7^ * V_ani_ ^2^ – 1.29*10^-7^ * V_ani_ + 1.65*10^-8^ (R^2^ = 0.99). For a 5:1 conductivity ratio, the relationship was C_ani_ = 4.02*10^-7^ * V_ani_ ^2^ – 8.51*10^-8^ * V_ani_ + 1.15*10^-8^ (R^2^ = 1.00). For a 7:1 conductivity ratio, the relationship was C_ani_ = 3.96*10^-7^ * V_ani_ ^2^ – 7.13*10^-8^ * V_ani_ + 1.02*10^-8^ (R^2^ = 1.00). Finally, for a 10:1 conductivity ratio, the relationship was C_ani_ = 3.95*10^-7^ V_ani_ ^2^ – 6.22*10^-8^ * V_ani_ + 9.35*10^-9^ (R^2^ = 1.00). (b) Similarly, the relationship between isotropic conduction velocity and anisotropic conductivity was studied for each of the four different conductivity ratio conditions. For a 3:1 anisotropic to isotropic conductivity ratio, the relationship between isotropic conduction velocit (V_iso_) and anisotropic conductivity (C_ani_) was C_ani_ = 1.92*10^-6^ * V_iso_ ^2^ – 3.77*10^-7^ * V_iso_ + 2.33*10^-8^ (R^2^ = 0.99). For a 5:1 conductivity ratio, the relationship was C_ani_ = 2.16*10^-6^ * V_iso_ ^2^ – 3.10*10^-7^ * V_iso_ + 1.73*10^-8^ (R^2^ = 0.99). For a 7:1 conductivity ratio, the relationship was C_ani_ = 2.29*10^-6^ * V_iso_ ^2^ – 2.58*10^-7^ * V_iso_ + 1.38*10^-8^ (R^2^ = 0.99). Finally, for a 10:1 conductivity ratio, the relationship was C_ani_ = 2.48*10^-6^ * V_iso_ ^2^ – 2.24*10^-7^ * V_iso_ + 1.18*10^-8^ (R^2^ = 1.00).

### Fiber Orientation Selection

A total of nine studies characterizing cardiac fiber orientation using DT-MRI were reviewed with N=45 subjects.(9–12,33–37) All subjects included in this review were considered healthy, volunteers, or normal. All but two studies were in humans and five of the nine studies were *in vivo*. Age ranges were reported in two studies to be 26 to 41 and 17 to 74 years.(10,12) For the five studies that reported sex, 20 subjects were male, and 12 were female.(10,12,34–36) Two papers reported helix angle data for fewer than the total number of participants in their respective studies. Reese, et al. reported helix angle data for two subjects while four were studied.(37) Tseng, et al. reported helix angle data in figure format for four of five subjects. This small degree of attrition is accounted for in the total number of 45 participants cited above.(10) Epicardial and endocardial fiber orientation are considered from each study and presented in **Fig. 3 (a-b)**. The range of helix angles 40° to 80° was selected for endocardial fiber orientations and is highlighted in **Fig. 3(a)**. Similarly, the range of helix angles -80° to -40° was selected for epicardial fiber orientations and is highlighted in **Fig. 3(b)**. The resulting five fiber orientations are shown in **Fig. 3(c)**.

**Fig 3.**
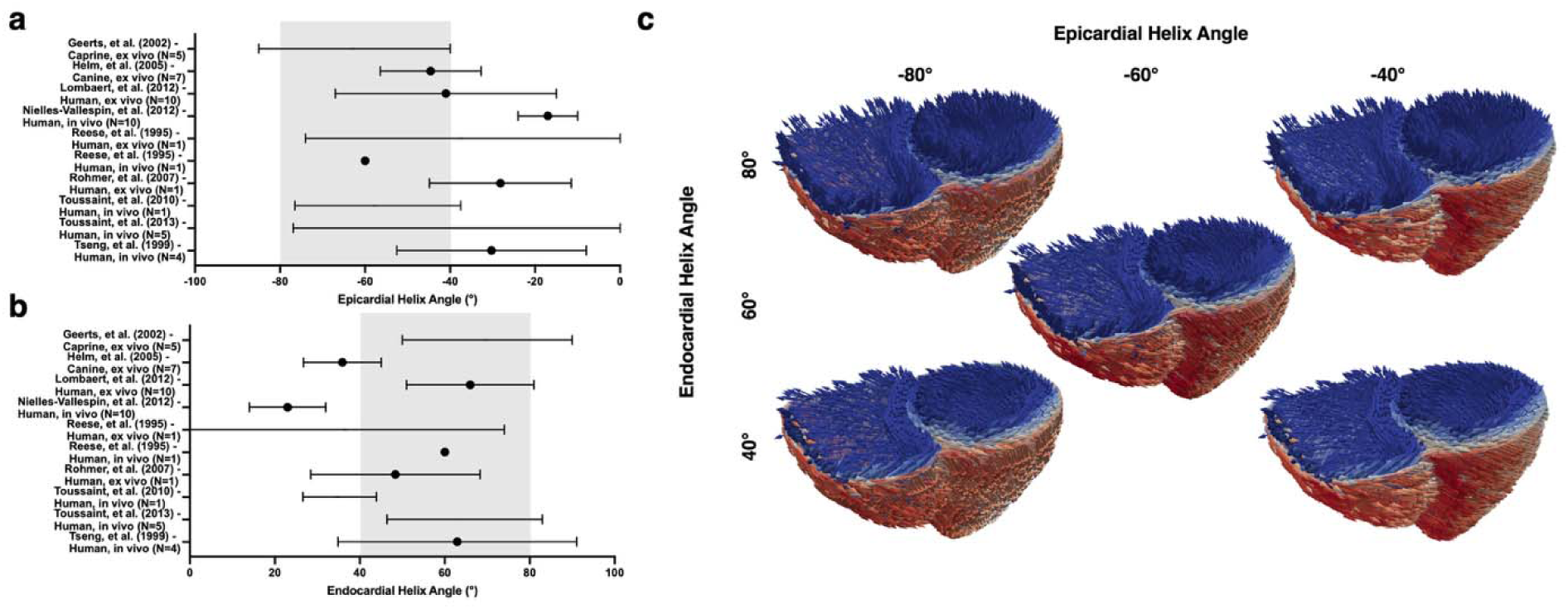
(a) Nine studies using cardiac DT-MRI to characterize fiber orientation in a total of forty-five subjects were reviewed. For each study, epicardial helix angle is represented. Points denote averages, error bars without points denote ranges, and error bars with points denote averages with standard deviation. The highlighted -80° to -40° range was selected as the parameter space of interest in this study. (b) Endocardial helix angles from the literature are similarly represented. The highlighted 40° to 80° range was selected as the parameter space of interest in this study. (c) Five meshes with variable endocardial and epicardial helix angles were generated using a rule-based algorithm for assigning myocardial fiber orientation.

### Electrocardiogram Definition and Pacing Conditions

Electrocardiograms were successfully generated by placing virtual leads in the usual locations for limb leads and along the precordium (**Fig. 1(a)**). Classic findings associated with LBBB and RBBB were replicated in this study (**Fig. 1(c)**). Notably, V1 was negative while V6 was positive in the late QRS for LBBB, while the opposite was observed for RBBB. A twelve-lead electrocardiogram is shown for an example site of ectopy in **Fig 1(d)**. Finally, ectopy location was observed to change QRS morphology considerably (**Fig. 1(e)**).

### Effect of Fiber Orientation on Depolarization Dynamics

For each of the eight pacing conditions and five fiber meshes, a twelve-lead EKG was generated. A representative sample of the electrocardiographic waveform data is summarized using Lead I in **Fig. 4**.

**Fig 4.**
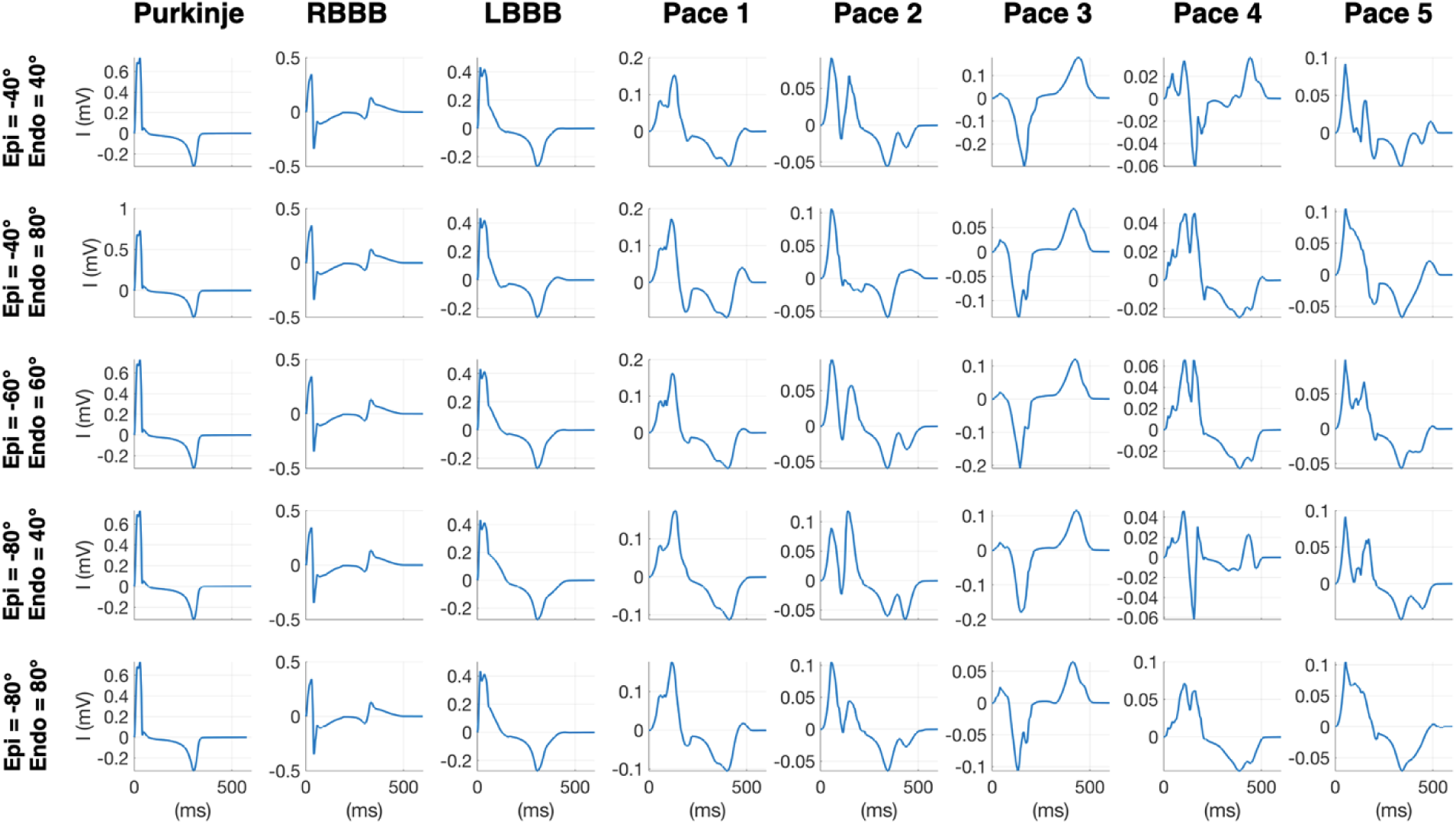
Lead I is shown for each experimental condition. Narrow QRS complexes are noted for the Purkinje control and bundle branch blocks, compared to the ectopy conditions. In the ectopy conditions, QRS morphology is noted to change considerably with varying fiber orientations.

Similarly, for each of the eight pacing conditions and five fiber meshes, activation maps were generated. **Fig. 5** demonstrates the subtle effect of altered fiber orientation on depolarization dynamics.

**Fig 5.**
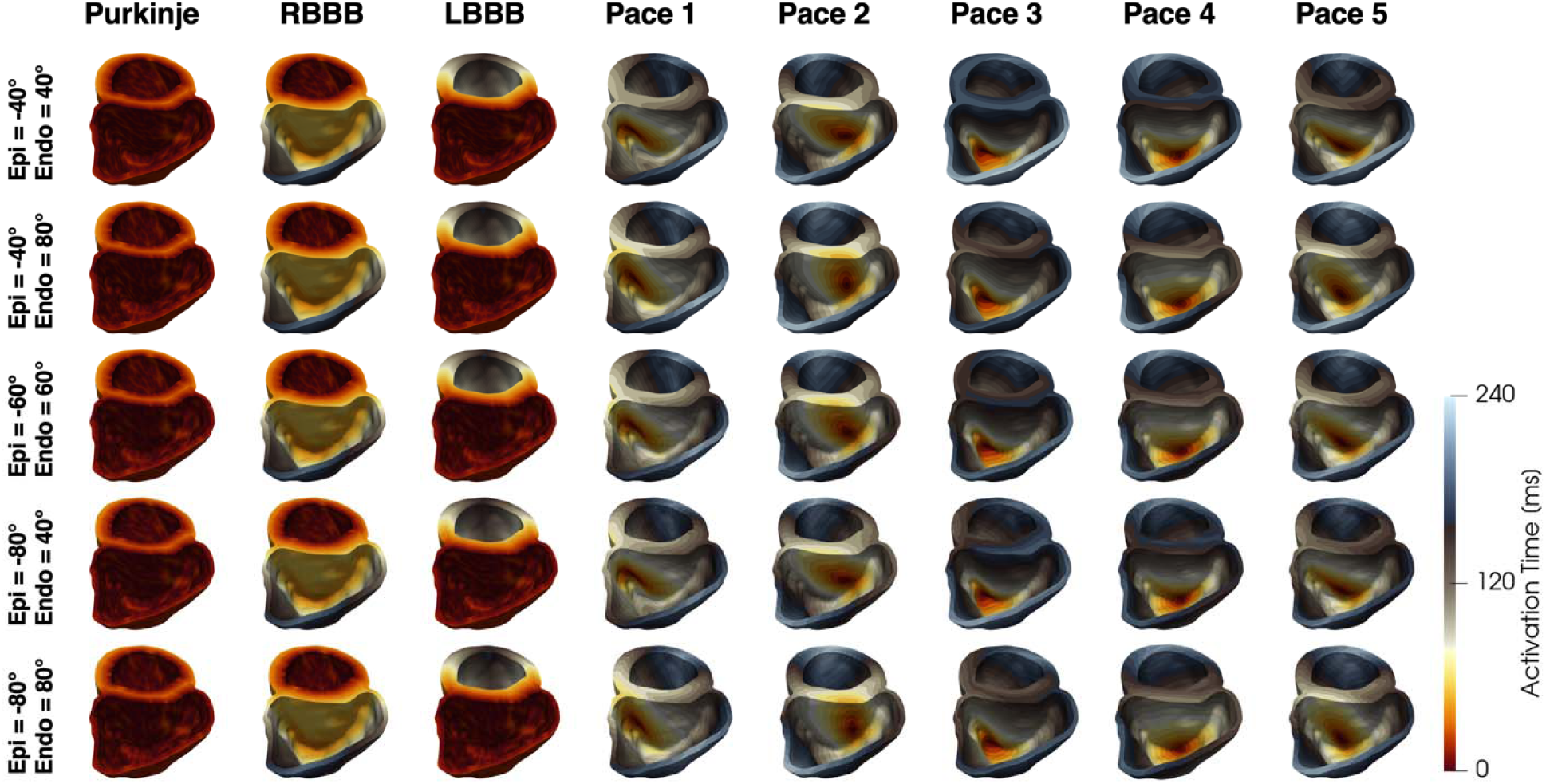
Activation maps are shown for each experimental condition. When modeling ectopy, fiber orientation is noted to subtly change the activation maps.

Finally, QRS duration was measured for each of the experimental conditions and is reported below in **Fig. 6**. **Fig. 6(a)** demonstrates the effect of altering fiber phenotype on QRS duration considering normal conduction and bundle branch block. **Fig. 6(b)** demonstrates the effect of altered fiber phenotype on QRS duration in each of the five ectopy simulations.

**Fig 6.**
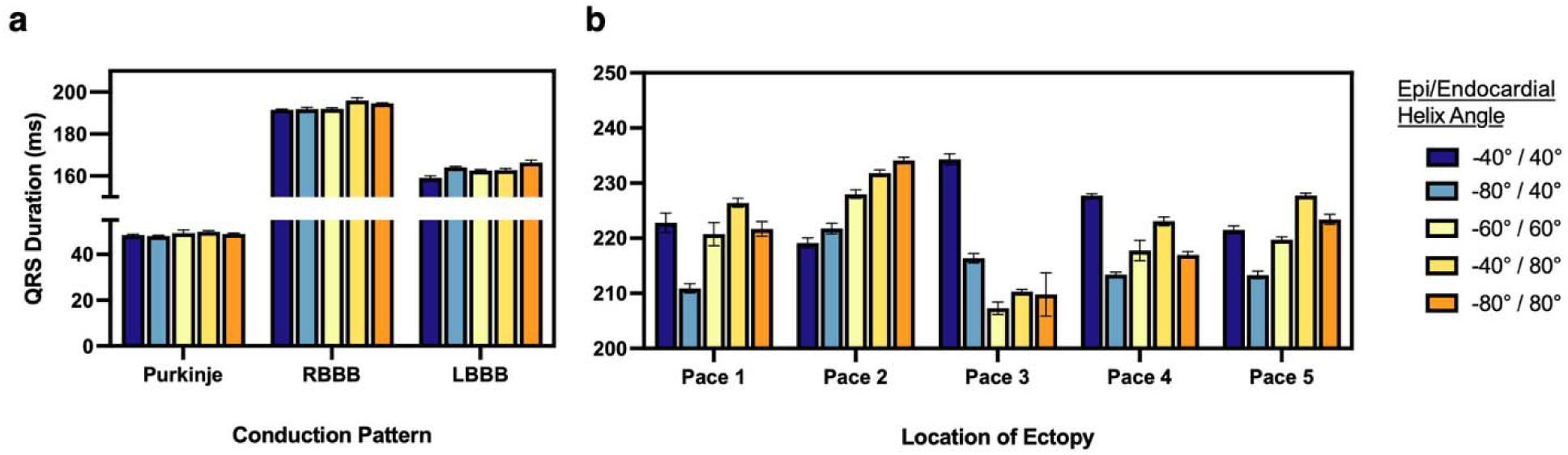
(a) QRS duration is measured for Purkinje, RBBB, and LBBB cases for all fiber orientations. An ANOVA considering variance in each of the three pacing groups was significant to *p*<0.01. Variations in fiber orientation led to a 1.88, 4.49, and 7.48 ms (3.92, 2.35, and 4.70%) increase to QRS duration in the Purkinje, RBBB, and LBBB pacing conditions. (b) A similar analysis was performed considering fiber orientations and each of the five ectopy locations. An ANOVA considering the variance in each of the five pacing conditions was significant to *p*<0.01. Variations in fiber orientation led to a 15.51, 14.97, 27.00, 14.33, and 14.40 ms (7.36, 6.83, 13.02, 6.72, and 6.75%) prolongation to QRS duration in pacing locations one through five, respectively.

In the case of normal Purkinje, RBBB, and LBBB conduction, changes to fiber orientation had a modest effect on QRS duration. An ANOVA considering variance for each of the three pacing conditions was significant to *p*<0.01. For normal Purkinje network conduction, altering fiber orientation was associated with a 3.92% increase in QRS duration when comparing the widest and narrowest QRS intervals for all conditions. For RBBB, altered fiber orientation accounted for a 2.35% increase to QRS duration. Finally, for LBBB, altered fiber orientation accounted for a 4.70% increase to QRS duration. **Table 1** summarizes the effect of fiber phenotype on QRS duration in native conduction, RBBB, and LBBB.

**Table 1.** Effect of Fiber Phenotype on QRS Prolongation in Control Conditions.

| Condition | Fiber Phenotype (Epi / Endo) | QRS Duration | Effect Size (ms (%)) | $p$-value |
| --- | --- | --- | --- | --- |
| Purkinje Conduction |  |  |  |  |
| Shortest QRS | -80° / 40° | 48.04 ± 0.33 ms | 1.88 ms (3.92%) | $p<0.01$ |
| Longest QRS | -40° / 80° | 49.92 ± 0.43 ms |  |  |
| RBBB |  |  |  |  |
| Shortest QRS | -40° / 40° | 191.53 ± 0.37 ms | 4.49 ms (2.35%) | $p<0.01$ |
| Longest QRS | -40° / 80° | 196.02 ± 1.24 ms |  |  |
| LBBB |  |  |  |  |
| Shortest QRS | -40° / 40° | 159.00 ± 1.09 ms | 7.48 ms (4.70%) | $p<0.01$ |
| Longest QRS | -80° / 80° | 166.48 ± 1.06 ms |  |  |

In addition, for each of the five pacing locations mimicking ventricular ectopy, the effect of variable fiber phenotype was examined. An ANOVA considering variance for each of the five pacing conditions was significant to *p*<0.01. For each of the pacing conditions, changing fiber phenotype was associated with an increase in the QRS duration of between 14.33 ms (6.72 %) and 27.00 ms (13.02 %). **Table 2** summarizes the results of the fiber phenotype pacing simulations.

**Table 2.**
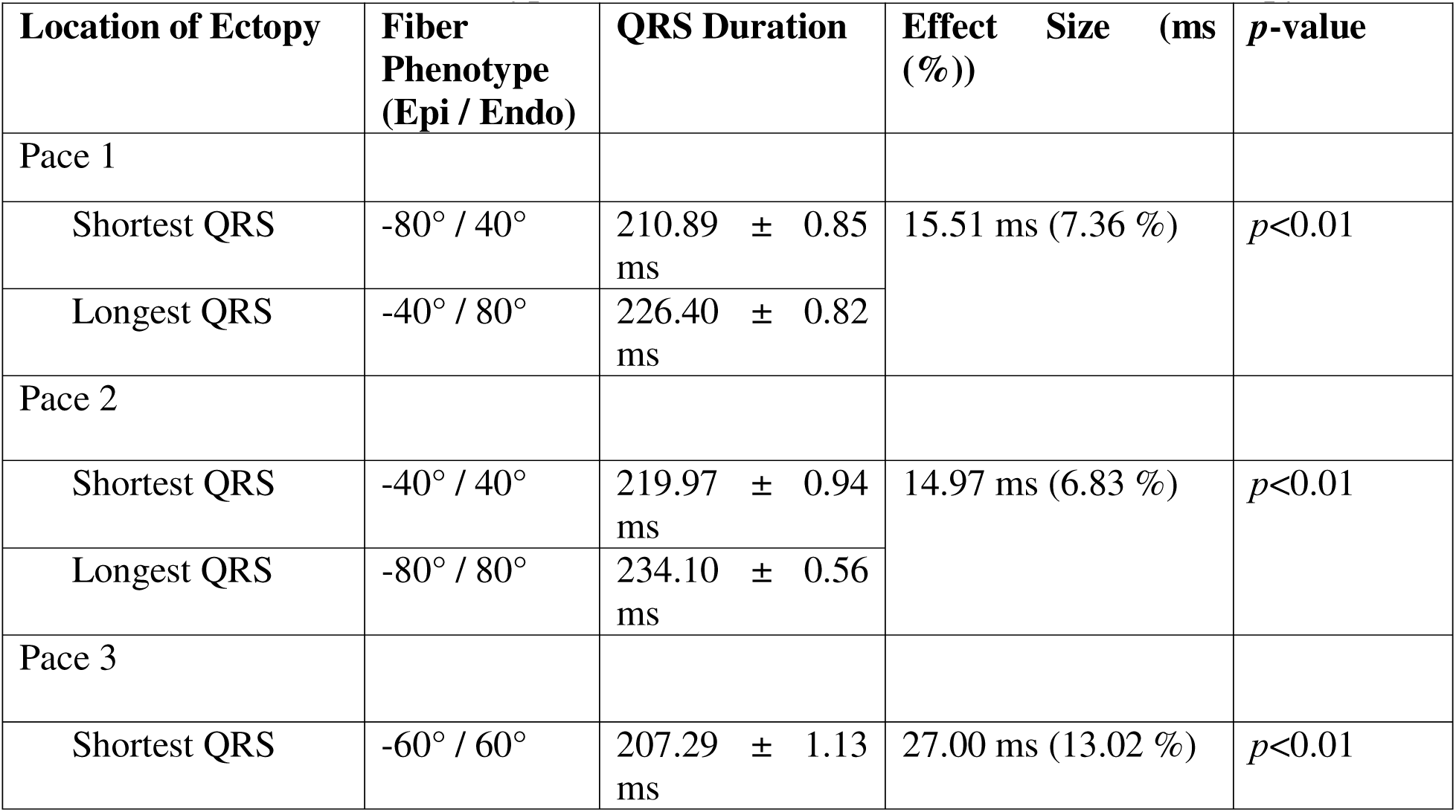
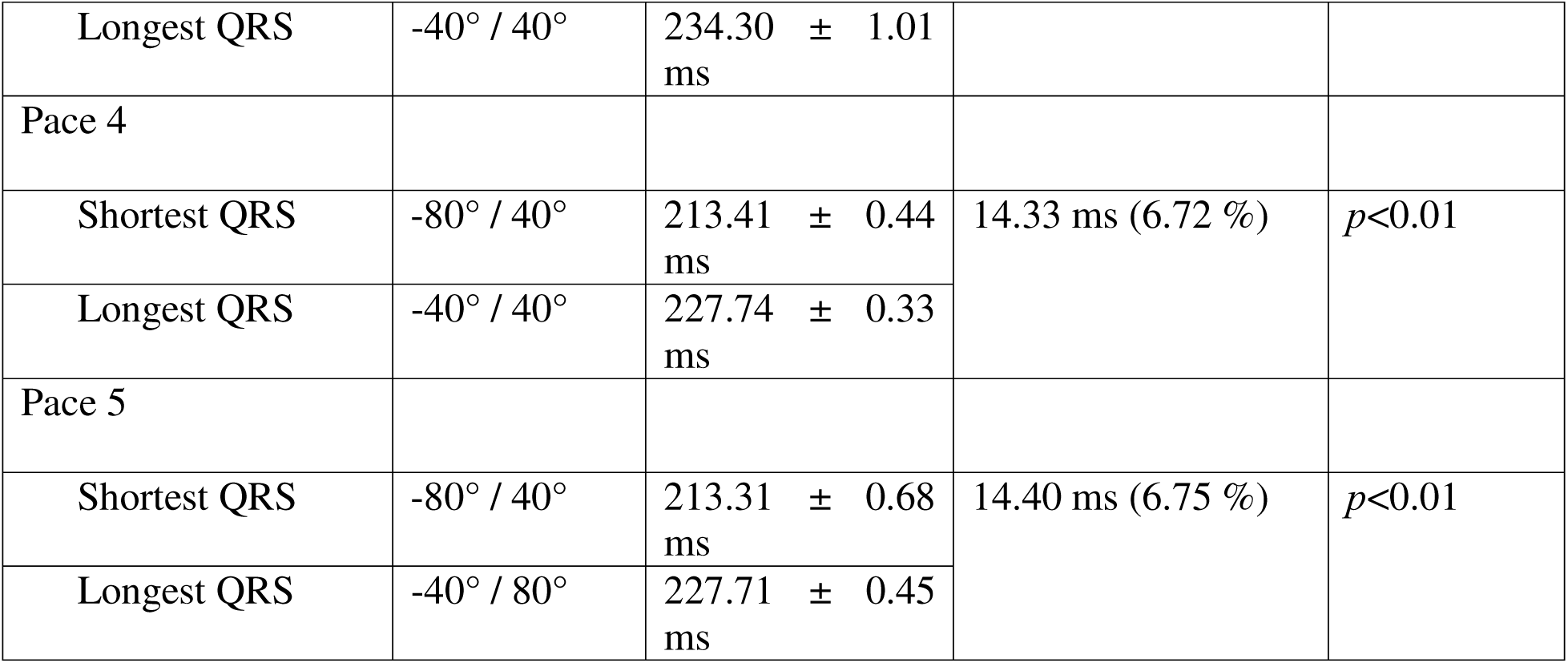
Effect of Fiber Phenotype on QRS Duration for Variable Sites of Ectopy.

| Location of Ectopy | Fiber Phenotype (Epi / Endo) | QRS Duration | Effect Size (ms (%)) | p-value |
| --- | --- | --- | --- | --- |
| Pace 1 |  |  |  |  |
| Shortest QRS | -80° / 40° | 210.89 ± 0.85 ms | 15.51 ms (7.36 %) | $p<0.01$ |
| Longest QRS | -40° / 80° | 226.40 ± 0.82 ms |  |  |
| Pace 2 |  |  |  |  |
| Shortest QRS | -40° / 40° | 219.97 ± 0.94 ms | 14.97 ms (6.83 %) | $p<0.01$ |
| Longest QRS | -80° / 80° | 234.10 ± 0.56 ms |  |  |
| Pace 3 |  |  |  |  |
| Shortest QRS | -60° / 60° | 207.29 ± 1.13 ms | 27.00 ms (13.02 %) | $p<0.01$ |
| Longest QRS | -40° / 40° | 234.30 ± 1.01<br>ms |  |  |
| Pace 4 |  |  |  |  |
| Shortest QRS | -80° / 40° | 213.41 ± 0.44<br>ms | 14.33 ms (6.72 %) | *p*<0.01 |
| Longest QRS | -40° / 40° | 227.74 ± 0.33<br>ms |  |  |
| Pace 5 |  |  |  |  |
| Shortest QRS | -80° / 40° | 213.31 ± 0.68<br>ms | 14.40 ms (6.75 %) | *p*<0.01 |
| Longest QRS | -40° / 80° | 227.71 ± 0.45<br>ms |  |  |

## DISCUSSION

It is currently unknown why 30 to 40% of patients with greater than 10% PVC burden and 13.8% of patients with single lead or dual chamber pacemakers develop cardiomyopathy (2,7). In both PVC and right ventricular pacing, electrical dyssynchrony evidenced by prolonged QRS and mechanical dyssynchrony seen on echocardiogram are known to be poor prognostic signs associated with increased risk of developing cardiomyopathy. In this work, we use finite element modeling to examine the effect of altered fiber orientation on QRS duration. Our results clinically implicate knowledge of patient-specific fiber orientation in both prognosis of bundle branch block and PVCs as well as in preclinical planning for patients undergoing right ventricular pacing.

In the first aim of this study, we hypothesized that a cardiac slab of well-defined dimensions would allow for tuning of anisotropic and isotropic conductivities to obtain desired conduction velocities. We used the well-studied geometry proposed by Niederer et al. (2011) (38). By tuning anisotropic and isotropic conductivities and measuring the resulting conduction velocities, our team obtained an anisotropic conduction velocity of 0.49 m/s and an isotropic conduction velocity of 0.21 m/s. This is similar to what has been documented *in vivo* for human myocardium of 0.5 meters per second and 0.17 m/s in the anisotropic and isotropic directions, respectively (32). The relationship between conductivity and conduction velocity was also found to be parabolic in the study, in agreement with what has been previously reported (39).

In the second aim of this study, a review of the DT-MRI literature was performed to describe the population variation in epicardial and endocardial helix angles. Despite forty-five subjects from nine studies all being healthy, there was a striking degree of epicardial and endocardial helix angle variation. Epicardial helix angle ranged between -76.9° and 0°, and endocardial helix angle ranged from 0° to 90°.(9–12,33–37) Fiber orientations in a structurally abnormal heart would be expected to exhibit an even broader spread of plausible helix angles. Sex was reported for only three studies with human subjects, ranging between 20 and 60% female. For these three studies, sex differences in predominant fiber phenotype were not observed.(10–12) Finally, only two studies reported the ages of subjects. Tseng, et al (1999)’s study population of five subjects ranged between 26 and 41 years old, while Lombaert et al (2011)’s sample of ten subjects ranged between 17 and 74 years old.(10,12) Both studies exhibited similar mean and standard deviation helix angles, despite Lombaert’s sample having both older and younger subjects. Taken together, the contribution of age and sex on variations to epicardial and endocardial helix angle as measured by DT-MRI in healthy subjects appears to be somewhat limited compared to the overall high level of heterogeneity in the population.

The results of this descriptive analysis were then used to generate five cardiac fiber phenotypes based on clinically relevant epicardial and endocardial helix angles. Much of the existing computational literature uses static values of epicardial and endocardial helix angles of - 60° and 60°, respectively. This study is unique in its characterization of the parameter space using a range of plausible fiber orientations based on the reported helix angles in the DT-MRI literature.

In the third aim of this study, we replicated classic electrocardiogram findings with various pacing conditions in our model. First, in normal Purkinje-mediated conduction, a narrow QRS was replicated. As expected, for RBBB, the late QRS was positive in V1 and negative in V6, while the opposite was observed for LBBB **(Fig. 1(c)).** Right ventricular septal ectopy resulted in a wide QRS and negative deflection in EKG leads II, III, and aVF (**Fig. 1 (d)**).

Importantly, our study shows that the alteration of fiber orientation can prolong the QRS to a clinically significant degree. Broadly, widened QRS complexes have been shown to be associated with impaired systolic function(40–42). Specifically in the setting of LBBB, the REVERSE trial found that in 369 patients, for every 10 ms increase to QRS duration on baseline EKG, there was a corresponding 5.7 mL/m^2^ decrease to left ventricular end systolic volume index (LVESVi) with initiation of cardiac resynchronization therapy (CRT) (43). Considering the 7.48 ms prolongation of QRS with altered fiber orientation in this study’s model of LBBB, it is possible that fiber orientation could be associated with a clinically significant decrease to LVESVi in patients with LBBB, and therefore decreased ejection fraction (43,44).

Similarly, prior studies have noted the relationship between prolonged QRS in patients with frequent PVCs and increased risk of developing cardiomyopathy. Deyell, et al. (2012) reported that in 114 patients with ≥ 10% PVC burden, for every 10 ms prolongation in QRS duration of ectopic beats, there was a 12.09 adjusted odds ratio for developing left ventricular dysfunction (5). Yokokawa et al. (2012) found that in 294 patients with idiopathic PVCs, QRS duration greater than 150 ms differentiated patients with and without cardiomyopathy with sensitivity of 0.80, specificity of 0.52, and AUC of 0.66 (4). In this paper we show a 14.33 to 27.00 ms (6.72 to 13.02%) prolongation in QRS duration with altered fiber phenotype, suggesting that fiber orientation may play an important role in potentiating risk of developing cardiomyopathy in patients with frequent PVCs.

Importantly, increased QRS duration is also known to confer greater risk of developing cardiomyopathy in patients undergoing RV pacing. In a retrospective study of 1,750 patients, Khurshid, et al. (2016) showed that for every 10 ms prolongation of QRS duration, there was an adjusted odds ratio of 1.34 for presence of PICM (45). For QRS interval duration greater than 150 ms, sensitivity for PICM was 95%. Considering the 14.33 to 27.00 ms prolongation to QRS noted with altered fiber orientation in this study, there may be a role for pre-operative quantification of fiber orientation to assist in optimal lead placement. Notably in this study, we also show that among five candidate pacing locations the most and least optimal pacing sites as measured by QRS duration depend heavily on the fiber network considered. For example, in the case of a fiber orientation with epicardial and endocardial helix angles of -40° and 40°, respectively, we note that Pace 2 resulted in the shortest QRS interval, while Pace 3 resulted in the longest QRS interval. Considering a fiber network with the same epicardial helix angle but steeper endocardial helix angle (epicardial and endocardial helix angles of -40° and 80°, respectively), we note the most and least optimal pacing sites switch positions, with Pace 2 resulting in the longest QRS interval and Pace 3 resulting in the shortest. This suggests that patient-specific knowledge of fiber orientation may be beneficial in preoperative planning for optimized lead placement, and therefore reduce the overall risk of developing PICM.

## LIMITATIONS

This study has several limitations. First, our methods are entirely computational. There is therefore uncertainty around the clinical correlation between the simulated pacing sites and shortened QRS intervals *in vivo*. However, every effort was made to ensure the clinical relevance of our model. In particular, we make use of a well described benchmark, and tune conduction velocities to values observed *in vivo*. Future work will evaluate the feasibility of preoperative fiber orientation mapping in a porcine model. In addition, future clinical work will examine patients with frequent PVCs, with and without cardiomyopathy, and study the relationship between measured fiber orientation using DT-MRI and location of ectopy. Among patients with frequent PVCs originating from the same region of the heart, epicardial and endocardial fiber orientation would be expected to differ for patients with and without cardiomyopathy.

Second, this study did not account for electromechanical coupling. Future work will evaluate the effect of fiber orientation on mechanical dyssynchrony.

Third, this work did not examine the effect of variable anisotropic to isotropic conduction ratios. The expected effect of altering this variable would be to observe more dramatic changes in QRS duration for greater degrees of anisotropic conduction. Future work will challenge the results of this work through a sensitivity analysis considering variable anisotropic to isotropic conduction ratios.

Fourth, fiber orientations as measured by DT-MRI vary significantly *in vivo*. On the one hand, significant population variability of fiber phenotype is a significant motivational factor for the presented research, since it could plausibly account for some degree of unfavorable electromechanics. However, it is unlikely that each phenotype studied in this work is equally likely to occur clinically. Future work should identify the source of variation in fiber orientation between subjects by performing subgroup analyses in larger populations. These would include age, sex, and medical comorbidities, details often absent from the cited literature above.

Finally, the subjects that informed selection for relevant helix angles in this work—as well as the subject used to generate the biventricular model used in this study—were all healthy. Patients with frequent PVCs and patients who undergo RV pacing often have comorbid structural heart disease. Those with an underlying cardiomyopathy, frequent PVCs, and increased pacing burden may therefore present with a mixed phenotype. In addition, patients with underlying cardiomyopathy often exhibit cardiac fibrosis, which would significantly alter the conduction velocities involved in affected areas. In the presented work, we did not model the effect of fibrosis. The goal of this work was to make conclusions to the possible contribution of fiber phenotype, so the inclusion of cardiac fibrosis in biventricular electrophysical modeling was reserved for future work.

## CONCLUSIONS

In this study, we examine the impact of fiber orientation on prolonged QRS duration in the setting of PICM and PVC myopathy. Our findings implicate fiber orientation as an important risk potentiator and suggest the possible future role of fiber orientation quantification and electrophysiological modeling preoperatively for optimal lead location selection in patients undergoing pacemaker placement. Future work will examine the feasibility and accuracy of such modeling in animal models of ventricular ectopy, and in observational clinical work.

## Data Availability

All data produced in the present study are available upon reasonable request to the authors.

## ACKNOWLEDGEMENTS

SP was a recipient of the Sarnoff Cardiovascular research fellowship and the Jacob P. Deerhake Research Award through the University of Michigan’s Department of Internal Medicine. Our thanks to Dr. Ian Chen MD PhD at the Veterans Affairs Hospital in Palo Alto, CA, who provided us with the biventricular model used in this study.

## Notes

### Competing Interest Statement

The authors have declared no competing interest.

### Author Declarations

IRB of Stanford University gave ethical approval for this work (IRB#39377).

